# Artificial neural networks to predict virological and immunological success in HIV patients under antiretroviral therapy from a nationwide cohort in Colombia, using the SISCAC database

**DOI:** 10.1101/2024.10.26.24316181

**Authors:** Alberto Buitrago-Gutierrez, Alexandra Porras-Ramirez

## Abstract

**Objective:** This study aimed to develop predictive models both for viral suppression and immunological reconstitution using a standard set of reported variables in a nationwide database system (SISCAC) from a cohort of patients living with HIV in Colombia.

**Materials and Methods:** We included 2.182 patients with no missing data related to the outcomes of interest, during a 12 month follow up period. We randomly assigned a 0,7 proportion of this cohort to de training dataset for 2 different predictive models (logistic regression, artificial neural networks). The AUC/ROC results were compared with those obtained through the construction of artificial neural networks with the specified parameters.

**Results:** From a cohort of 2182 patients, 85,79% were male and at HIV diagnosis, the mean value of the CD4 count was 342 x mm3. The logistic regression models obtained AUC/ROC accuracy for the outcomes “suppressed viral load” 0,7, “undetectable viral load” of 0,66 and “immunological reconstitution” 0,83; whereas the artificial neural network perceptron multilayer obtained AUC/ROC of 0,77, 0.69 and 0,87 for the same outcomes.

**Conclusions:** The selection of specific variables from a nationwide database in Colombia with quality control purposes allowed us to generate predictive models with an initial evaluation of performance regarding three predefined outcomes for virological and immunological success.

## Introduction

Human immunodeficiency virus (HIV) infection and AIDS continue to generate a significant burden of morbidity and mortality worldwide, with developing countries contributing the most cases.

At a global level, the strategy of the Joint United Nations Program on HIV and AIDS (UNAIDS) was generated to direct and prioritize efforts to control the disease. Thus, for 2020, the goal of 90-90-90 was proposed, which refers to achieving a diagnosis of 90% of people living with HIV, ensuring access to antiretroviral therapy for 90% of those diagnosed and achieving at least 90% of virologic suppression in those on treatment. Additionally, and more recently, that goal was revised to bring it to a goal of 95% in a horizon set until the year 2030 (1).

In Colombia exists a database in the high cost account (“Cuenta de alto costo”, in spanish), this institution has the task of monitoring high cost diseases under the regulations defined by the Ministry of Health and Social Protection of Colombia, it thereby reports the data generated by the different health insurers and providers of the country’s social security system, including those referring to the care of people living with HIV (PLWH) in the country; latest results showing an increase in the number of annual cases from 35,000 in 2012 to 123,490 cases by 2020 (1).

It is essential to introduce technological and computational tools in the clinical environment and with data obtained from our own population of patients evaluate classification and prediction models that can support the clinical approach; in our country the national SISCAC system can provide a robust database to build classification and prediction models that facilitate these clinical decisions and the creation of data driven algorithms and protocols. Proposals for the use of machine learning in health care are supported with several goals, such as enhancing the patient experience, optimizing population health, reducing costs, and improving the working conditions of health workers. (2–15)

Studies such as that of Robbins GK et al, through the collection of data from electronic medical record records in two tertiary care centers in Boston (United States); the Massachusetts General Hospital and the Brigham and Women’s Hospital, evaluated through a multivariable logistic model which variables were associated with a greater risk of virological failure in patients who entered care of the HIV clinic within the first year from the start of care and treatment (10).

Wang D. et al. developed artificial neural network models to predict the response to therapy using data from resistance genotypes and clinical information, they compared the results obtained through the artificial neural network model with other methodologies such as random forests and support vector machine algorithms. Using data from 1,204 episodes of treatment changes for antiretroviral therapy, obtained from the RDI (HIV Resistance Response Database Initiative) database, they sought to predict the behavior of viral load in the different models, observing a similar correlation between the RF and SVM models with the artificial neural network (17).

The results showed r^2^ correlations in the predicted viral load with the observed value that varied between 0.318-0.546 for the neural network, 0.590-0.751 for the random forests model and 0.300 to 0.720 for the support vector machine model. Subsequently, by combining the outputs of the 3 models, improvements were achieved in the correlations of the virological predictions obtained and those observed. The authors concluded that the introduced models (RF’s and SVM) demonstrated similar performance to the neural network but that the correlation results could improve with their combined application. (17).

Neural networks use a structure of neurons or nodes to analyze the interactions between a group of covariates to predict an outcome; these neurons are organized by layers that are associated in an input layer, intermediate hidden layers, and an output layer. The weighted sum of neural inputs is somewhat analogous to the coefficients in linear and logistic regressions, and the connection weights are iteratively adjusted by the learning algorithm to minimize error and improve predictions (18). It is considered a strength for neural networks its ability to fit interactions between variables in non-linear associations without any prior user specification (11,19).

Our objective was to evaluate models for the prediction of virological and immunological success with three predefined outcomes; viral load category “suppressed” or viral load <200 cop/mL, viral load category “undetectable” or viral load <20 cop/mL; and the immunological outcome considering a CD4 lymphocyte count at the end of the 12-month follow-up period above 350 cells x mm3 as indicative of “immunological reconstitution”. Using the clinical and laboratory variables obtained from the SISCAC registries for HIV/AIDS and with particular interest in evaluating the inclusion of an integrase inhibitor in the antiretroviral therapy. We sought to determine which model offers better performance through a comparison of a multivariable logistic regression model and artificial neural networks.

## Materials and Methods

This study pursued the training and validation of models using anonymous data from a cohort of patients from a specialized HIV care provider in Bogotá (Col), using the SISCAC database of the high-cost account. (https://cuentadealtocosto.org/site/general/comunicado-siscac/) (21–23).

The analysis included only data obtained from adult patients (18 years +) reported within the period of 12 months of follow-up between January 1^st^ 2022 and January 1^st^ 2023, the data was previously uploaded to the high-cost account following a structured set of 99 variables, of which the final analysis took 15, either in their primary registry or recoded to be included in the modelling.

This study was approved by the Ethics and Research committee of Vidamedical IPS on March 17^th^, 2023. All methods followed national (Resolution 8430 of 1993, stated by the Colombian Health Ministry) and international (The Declaration of Helsinki) standards. It was a secondary data use, observational and retrospective study; therefore it did not require informed consent from participants.

### Variables and definitions

The demographic variables from the SISCAC notification record included age, sex, weight and height, the diagnosis of pregnancy at the time of notification; the presence of comorbidities such as active tuberculosis, hepatitis B and C virus coinfection, chronic kidney disease, coronary disease and neoplasms not associated with AIDS.

Additionally, variables related to the characterization of HIV infection were taken, such as the years of diagnosis at the time of data notification, the mechanism of transmission of HIV infection, the stage at the time of diagnosis, the CD4 lymphocyte count and viral load at diagnosis and also at the time of initiation of antiretroviral therapy; also at the end of the 12-month follow-up period (January 2022 to January 2023). Changes in antiretroviral therapy, the presence of treatment failures, the number of infectious disease specialty consultations within the 12-month follow-up (>3 or <3 visits in the 12-month period) were recorded; the months of effective dispensing of antiretroviral treatment, the presence of an integrase inhibitor (raltegravir, elvitegravir, dolutegravir) was recorded in the initial scheme and the final follow-up scheme for the 12-month period.

The variables were included in the models in a dichotomous way to facilitate their analysis. For example, the measurement of the months of dispensing therapy in the last year was categorized into patients with compliance greater than 95% and those below this goal, considering 100% dispensing in the total 12 months of the year, The variable on frequency of consultation by the infectious disease specialty was dichotomized according to the distribution found in the observations in >3 or <3 visits (21).

### Data source

The source of data were the records of the variables from the notification to the SISCAC database, and we included those considered relevant to the purpose of this investigation. The information is previously verified in accordance with the instructions for reporting information to the high-cost account for HIV/AIDS, regulated under Resolution 273 of 2019 of the Ministry of Health and Social Protection of Colombia (23) (Chrome-extension://efaidnbmnnibpcajpcglclefindmkaj/https://www.minsalud.gov.co/sites/rid/Lists/Bibli otecaDigital/RIDE/DE/DIJ/resolucion-273-de-2019.pdf) (23)

The data from ninety-nine variables corresponding to the administrative follow-up of the SISCAC was initially extracted (except cut-off date/Var.98 and serial code/Var.99) for a cohort of 6526 patients, belonging to an HIV service care provider in Colombia (Vidamedical IPS, https://www.vidamedicalips.com/). Patients included came from three cities (Bucaramanga, Cúcuta and Bogotá).

We discarded the records corresponding to patients under 18 years of age and the observations or patients that had missing data regarding the outcomes of interest at diagnosis, initiation of antiretroviral treatment, and at the end of follow-up (last 12 months between January 2022 and January 2023). By filtering for lost data and recoding them, a total of fifteen variables were included in the definitive analysis (See figure No. 1 Patient selection flowchart).

**Figure 1.**
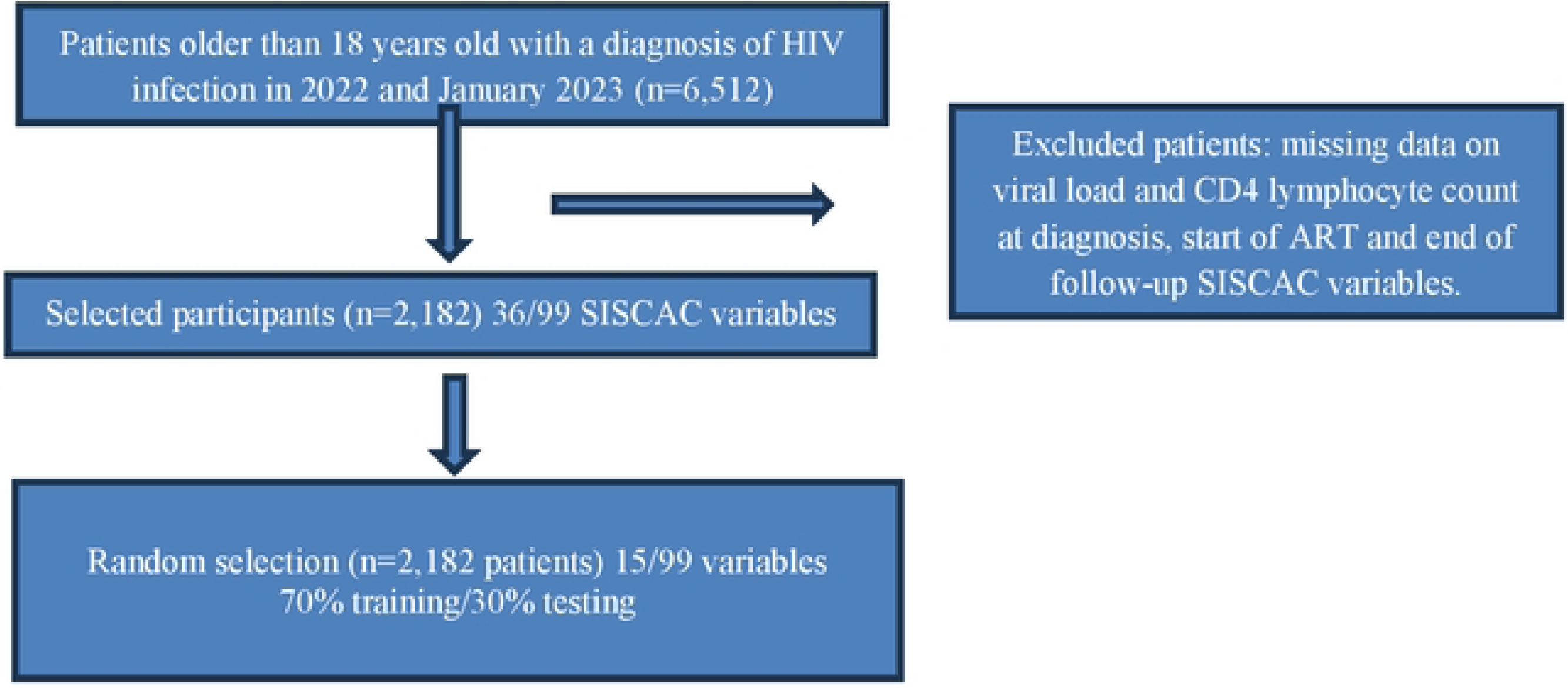
Population selection flowchart.

### Outcomes evaluated

We established three dependent variables or outcomes: suppressed viral load with a value <200 cop/ml, undetectable viral load with a value of <20 cop/ml and CD4 lymphocyte count >350 x mm3, all considered at the end of the twelve-month follow-up period. The CD4 count value was set arbitrarily by the investigators for the immunological reconstitution outcome, considering the mean value for the entire cohort in this parameter at the time of diagnosis.

### Ethical aspects

The study was performed through the secondary use of data previously collected in a periodically and regulated manner under resolution 273/2019 from the Ministry of Health and Social Protection of Colombia. (Chrome-extension://efaidnbmnnibpcajpcglclefindmkaj/https://www.minsalud.gov.co/sites/rid/Lists/Bibli otecaDigital/RIDE/DE/DIJ/resolucion-273-de-2019.pdf).

The study followed the recommendations for the secondary use of clinical data (24–26). The protocol was approved by the Vidamedical IPS research and ethics committee (https://www.vidamedicalips.com/). Data extraction was performed by computing personnel of the provider’s data analysis area and delivered to the researchers in the excel program format (Microsoft® Excel®).

To preserve the anonymity of the patients and since it was a secondary analysis of data already collected for the purposes of notification to the high-cost account (CAC), each patient had a natural consecutive number assigned, discarding the identification numbers and names.

### Statistical analysis

We performed a bivariate analysis to obtain crude association measures in the training or retrospective cohort, obtaining the respective OR’s (See tables 1, 2 and 3). The analysis then included the multivariate logistic regression model to adjust the variables for potential confusion and interaction, using the statistical program Stata Version 18.0 B. E, with the construction of the respective AUC//ROC curves for each of the outcomes; calculating in the multivariate model the O.R’s with the corresponding confidence intervals.

**Table 1.**
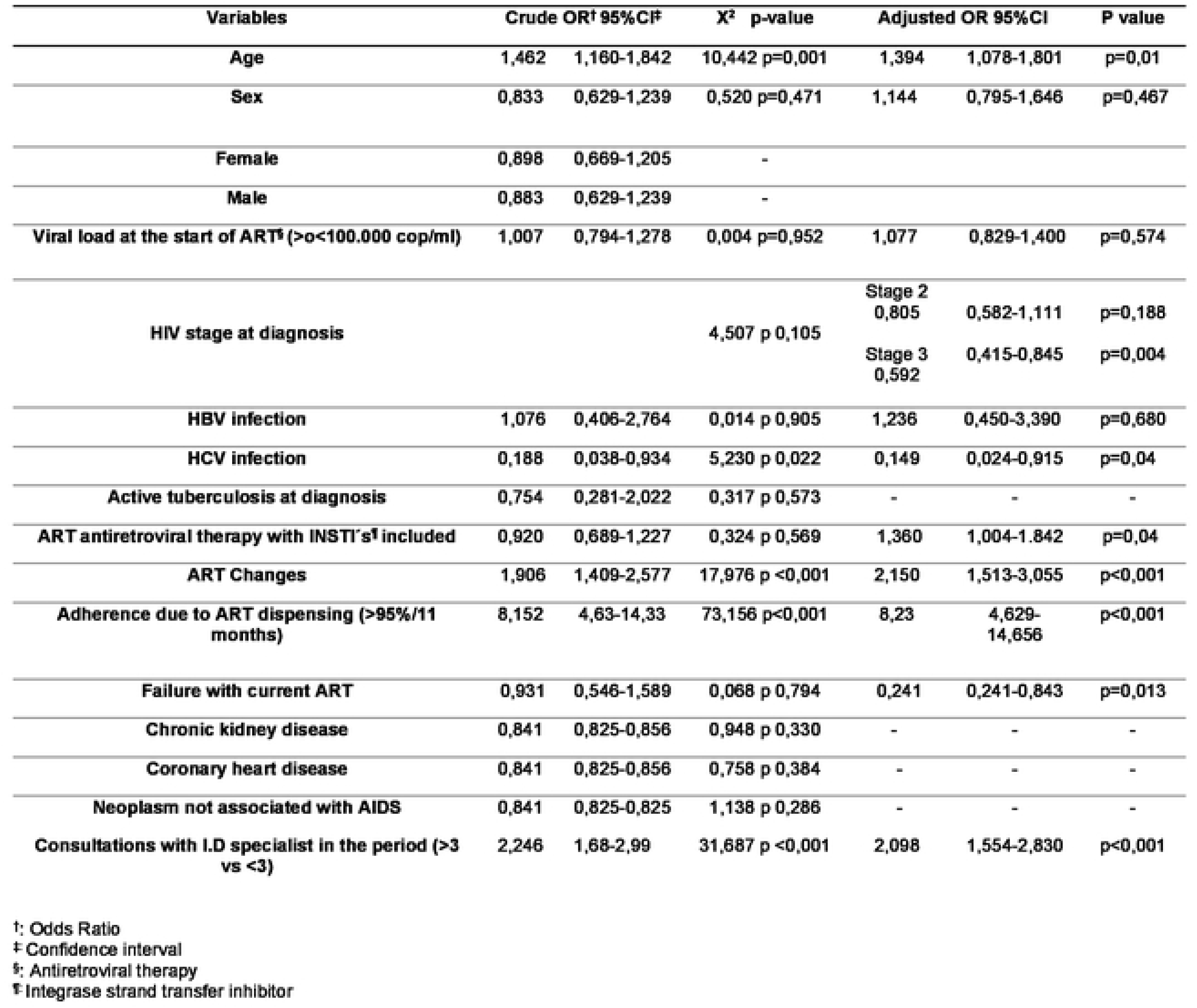
Bivariate and multivariate regression analysis for the outcome “suppressed viral load.”

| Variables | Crude OR<sup>†</sup> 95%CI<sup>‡</sup> |  | X<sup>2</sup> p-value | Adjusted OR 95%CI |  | P value |
| --- | --- | --- | --- | --- | --- | --- |
| Age | 1,462 | 1,160-1,842 | 10,442 p=0,001 | 1,394 | 1,078-1,801 | p=0,01 |
| Sex | 0,833 | 0,629-1,239 | 0,520 p=0,471 | 1,144 | 0,795-1,646 | p=0,467 |
| Female | 0,898 | 0,669-1,205 | - |  |  |  |
| Male | 0,883 | 0,629-1,239 | - |  |  |  |
| Viral load at the start of ART<sup>§</sup> (>0<100.000 cop/ml) | 1,007 | 0,794-1,278 | 0,004 p=0,952 | 1,077 | 0,829-1,400 | p=0,574 |
| HIV stage at diagnosis |  |  | 4,507 p 0,105 | Stage 2<br>0,805 | 0,582-1,111 | p=0,188 |
|  |  |  |  | Stage 3<br>0,592 | 0,415-0,845 | p=0,004 |
| HBV infection | 1,076 | 0,406-2,764 | 0,014 p 0,905 | 1,236 | 0,450-3,390 | p=0,680 |
| HCV infection | 0,188 | 0,038-0,934 | 5,230 p 0,022 | 0,149 | 0,024-0,915 | p=0,04 |
| Active tuberculosis at diagnosis | 0,754 | 0,281-2,022 | 0,317 p 0,573 | - | - | - |
| ART antiretroviral therapy with INSTI's<sup>¶</sup> included | 0,920 | 0,689-1,227 | 0,324 p 0,569 | 1,360 | 1,004-1,842 | p=0,04 |
| ART Changes | 1,906 | 1,409-2,577 | 17,976 p <0,001 | 2,150 | 1,513-3,055 | p<0,001 |
| Adherence due to ART dispensing (>95%/11 months) | 8,152 | 4,63-14,33 | 73,156 p<0,001 | 8,23 | 4,629-14,656 | p<0,001 |
| Failure with current ART | 0,931 | 0,546-1,589 | 0,068 p 0,794 | 0,241 | 0,241-0,843 | p=0,013 |
| Chronic kidney disease | 0,841 | 0,825-0,856 | 0,948 p 0,330 | - | - | - |
| Coronary heart disease | 0,841 | 0,825-0,856 | 0,758 p 0,384 | - | - | - |
| Neoplasm not associated with AIDS | 0,841 | 0,825-0,825 | 1,138 p 0,286 | - | - | - |
| Consultations with I.D specialist in the period (>3 vs <3) | 2,246 | 1,68-2,99 | 31,687 p <0,001 | 2,098 | 1,554-2,830 | p<0,001 |

†: Odds Ratio
‡ Confidence interval
§: Antiretroviral therapy
¶ Integrase strand transfer inhibitor

**Table 2.** Bivariate and multivariate regression analysis for the outcome “undetectable viral load”.

| Variables | Crude OR<sup>†</sup> 95%CI |  | X<sup>2</sup> p-value | Adjusted OR 95%CI |  | P value |
| --- | --- | --- | --- | --- | --- | --- |
| Age | 1,182 | 0,982-1,423 | 3,131 p=0,077 | 1,226 | 0,997-1,506 | p=0,05 |
| Sex | 0,999 | 0,766-1,302 | 0,000 p=0,991 | 1,191 | 0,896-1,583 | p=0,226 |
| Female | 0,999 | 0,795-1,254 | - |  |  |  |
| Male | 1,000 | 0,963-1,039 | - |  |  |  |
| Viral load at the start of ART<sup>§</sup> (>0<100.000 cop/ml) | 0,637 | 0,527-0,770 | 21,948 p<0,001 | 0,708 | 0,575-0,871 | p=0,001 |
| HIV stage at diagnosis |  |  | 28,926 p<0,001 | Stage2<br>0,783 | 0,602-1,019 | p=0,06 |
|  |  |  |  | Stage3<br>0,520 | 0,391-0,693 | p<0,001 |
| HBV infection | 1,076 | 0,498-2,329 | 0,035 p=0,852 | 1,186 | 0,524-2,686 | p=0,681 |
| HCV infection | 0,080 | 0,009-0,687 | 8,758 p=0,003 | 0,760 | 0,008-0,676 | p=0,021 |
| Active tuberculosis at diagnosis | 0,601 | 0,269-1,345 | 1,567 p=0,211 | - | - | - |
| ART antiretroviral therapy with INSTI's<sup>¶</sup> included | 0,851 | 0,675-1,075 | 1,835 p=0,176 | 1,154 | 0,901-1,477 | p=0,256 |
| ART Changes | 1,586 | 1,265-1,989 | 16,164 p<0,001 | 1,783 | 1,373-2,316 | p<0,001 |
| Adherence due to ART dispensing (>95%/11 months) | 2,618 | 1,991-3,443 | 50,036 p<0,001 | 2,854 | 2,135-3,815 | p<0,001 |
| Failure with current ART | 0,750 | 0,492-1,142 | 1,811 p=0,178 | 0,475 | 0,293-0,769 | p=0,002 |
| chronic kidney disease | 1,614 | 0,180-14,473 | 0,187 p=0,666 | - | - | - |
| Coronary heart disease | 0,712 | 0,693-0,731 | 1,616 p=0,204 | - | - | - |
| Neoplasm not associated with AIDS | 0,712 | 0,693-0,731 | 2,426 p=0,119 | - | - | - |
| Consultations with I.D specialist in the period (>3 vs <3)) | 1,648 | 1,277-2,128 | 14,904 p<0,001 | 1,505 | 1,154-1,965 | p=0,003 |

†: Odds Ratio
‡ Confidence interval
§: Antiretroviral therapy
¶ Integrase strand transfer inhibitor

**Table 3.** Bivariate and multivariate regression analysis for the outcome “immunological reconstitution”.

| Variables | Crude OR† 95%CI |  | X2 p-value | Adjusted OR 95%CI |  | p value |
| --- | --- | --- | --- | --- | --- | --- |
| Age | 0,634 | 0,527-0,762 | 23,605 p<0,001 | 0,986 | 0,778-1,250 | p=0,912 |
| Sex | 1,238 | 0,960-1,596 | 2,718 p=0,099 | 1,181 | 0,863-1,617 | p=0,297 |
| Female | 1,200 | 0,967-1,489 |  |  |  |  |
| Male | 0,969 | 0,932-1,007 |  |  |  |  |
| Viral load at the start of ART‡ (>0<100.000 cop/ml) | 0,582 | 0,483-0,702 | 32,678 p<0,001 | 1,402 | 1,107-1,776 | p=0,005 |
| HIV Stage at diagnosis |  |  | 556,626 p<0,001 | Stage 2<br>0,078<br>Stage 3<br>0,010 | 0,042-0,146<br>0,005-0,019 | p<0,001<br>p<0,001 |
| HBV infection | 1,174 | 0,543-2,540 | 0,167 p=0,683 | 0,935 | 0,360-2,428 | p=0,891 |
| HCV infection | 0,438 | 0,088-2,177 | 1,076 p=0,300 | 0,200 | 0,025-1,547 | p=0,123 |
| Active tuberculosis at diagnosis | 0,288 | 0,129-0,645 | 10,362 p=0,001 | - | - | - |
| ART antiretroviral therapy with INSTI's¶ included | 0,601 | 0,481-0,751 | 20,298 p<0,001 | 0,661 | 0,498-0,877 | p=0,004 |
| ART Changes | 1,794 | 1,432-2,249 | 26,203 p<0,001 | 2,594 | 1,927-3,492 | p<0,001 |
| Adherence due to ART dispensing (>95%/11 months) | 1,534 | 1,208-1,946 | 12,484 p<0,001 | 2,123 | 1,586-2,843 | p<0,001 |
| Failure with current ART | 0,749 | 0,495-1,135 | 1,865 p=0,172 | 0,471 | 0,277-0,801 | p=0,005 |
| Chronic kidney disease | 0,694 | 0,675-0,714 | 2,202 p=0,138 | - | - | - |
| Coronary heart disease | 0,439 | 0,062-3,120 | 0,717 p=0,397 | - | - | - |
| Neoplasm not associated with AIDS | 2,201 | 0,257-18,872 | 0,545 p=0,460 | - | - | - |
| Consultations with I.D specialist in the period (>3 vs <3)) | 1,539 | 1,194-1,984 | 11,203 p<0,001 | 1,392 | 1,017-1,905 | p=0,038 |

†: Odds Ratio
‡ Confidence interval
§: Antiretroviral therapy
¶ Integrase strand transfer inhibitor

For the creation of the neural network, we used IBM SPSS Statistics Version 28.0.0.0 program, which implements the “multilayer perceptron” network with a partition of 70% of the data in the training model and 30% in the testing; with a hyperbolic tangent hidden layer activation function and softmax output layer activation function (18)

In the construction of the artificial neural network we included ten factors given by the dichotomized predictive variables: age, sex, stage at diagnosis, start of antiretroviral therapy with any integrase inhibitor, coinfection with hepatitis B virus, coinfection with hepatitis C virus, active tuberculosis, changes in antiretroviral therapy, antiretroviral therapy failures, consultations by the I.D specialist >3 in the period of follow-up, chronic kidney disease, coronary disease, non-AIDS associated neoplasm and adherence to the antiretroviral therapy dispensing above 95%.

Additionally, as covariates we included viral load at the beginning of treatment and CD4 lymphocyte count at the beginning of treatment plus the value of the body mass index (BMI) (18).

The distribution of the factors and the quantitative covariates was done to maintain the assumption of independence, using the multilayer perceptron network, as a supervised learning technique, with feedback architecture, hyperbolic tangent activation function in the hidden layers and softmax in the hidden layer output, plus cross entropy error function (18).

## Results

From the 2182 patients included we identified a proportion of 85.79% for the male sex and 14.21% for the female sex. Regarding age, we observed for the entire cohort a mean of 33 years old (SD +/- 11.6 years), with an equal median, considering a normal distribution of this variable. For this cohort, the mean HIV diagnosis time was 3.3 years, with a minimum follow-up time from diagnosis of 3 months and a maximum of 18 years.

The reported mechanism of HIV transmission corresponded in 98.63% to sexual, with 1.37% grouping other different mechanisms and those not reported. At the time of diagnosis of HIV infection, the mean value of the CD4 lymphocyte count was 342 x mm3. The proportion of those coinfected with hepatitis B at the start of treatment was only 1.51%, while hepatitis C coinfection was lower with 0.27%. The proportion of patients coinfected with active tuberculosis was 1.15%.

During the initiation of antiretroviral therapy, the mean dispensing time of the antiretroviral therapy was 6.5 months (SD+/-4.45). The mean of therapy changes was 1.7 during de 12-month follow-up period, and the diagnosis of antiretroviral therapy failures was done on average 1.95 times with a minimum of one change and a maximum of three modifications in the antiretroviral therapy.

Regarding the outcome of “suppressed viral load” (<200 cop/ml), the following variables were observed as statistically significant: HIV stage at diagnosis (X2 4.507; p=0.105); hepatitis C virus infection at diagnosis (OR 0.188 95% CI 0.038-09.34; p=0.022); changes in antiretroviral therapy (OR 1.906 95% CI 1.409-2.577; p<0.001); adherence to antiretroviral therapy dispensing above 95% (OR 8.152 95%CI 4.63-14.33; p<0.001) and the record of >3 attentions for infectious diseases in the last year (OR 2.246 95%CI 1.68-2, 99; p<0.001) (See table 1). We used the bivariate and multivariate logistic regression analysis to generate the correspondent AUC/ROC curve for the outcome “suppressed viral load”, obtaining an area under de ROC curve of 0.7017.

When estimating the associations for the outcome variable “undetectable viral load” (<20 cop/ml) it was found that the variables: HIV stage at diagnosis (X2 28.926; p<0.001); viral load at the start of ART >100,000 cop/ml (OR 0.637 95% CI 0.527-0.770; p<0.001); hepatitis C virus infection at diagnosis (OR 0.08 95% CI 0.009-0.687; p 0.003); changes in antiretroviral therapy during the follow-up period (O,R 1.586 95% CI 1.265-1.989; p<0.001); adherence to the dispensing of antiretroviral therapy above 95% (OR 2.618 IC95% 1.991-3.443; p<0.001) and likewise the record of >3 attentions for the I.D specialist in the follow-up period (OR 1.648 IC95% 1.277-2.128; p<0.001) were statistically significant (See table 2).

We evaluated the accuracy for the outcome of “undetectable viral load” for the multivariable logistic regression model, with the variables previously listed, obtaining an AUC/ROC of 0,6605. (See figure 2).

**Figure 2.**
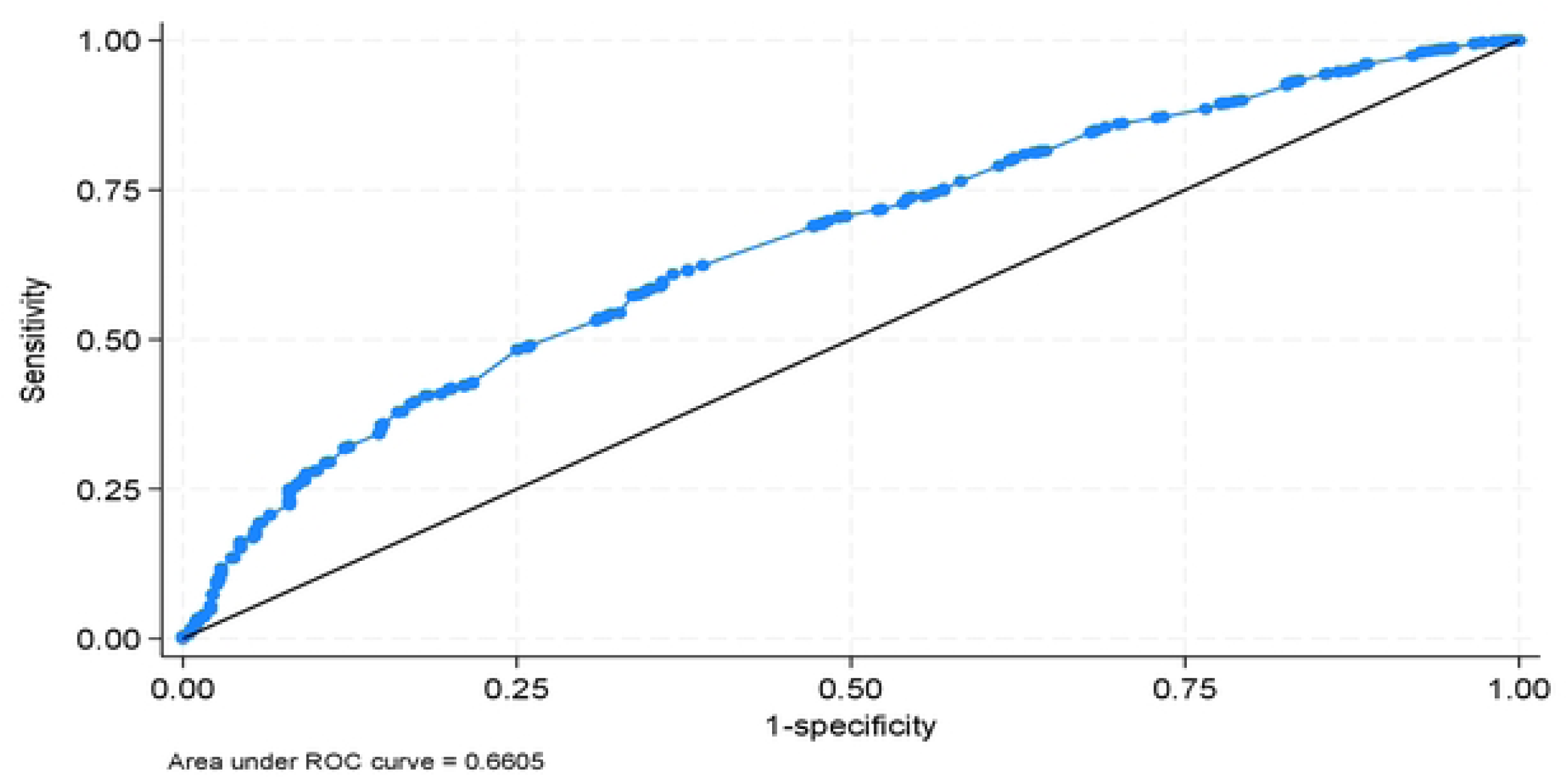
AUC/ROC for “undetectable viral load” multivariable logistic regression.

Finally we obtained crude and adjusted O.R’s for the outcome immunological reconstitution at the end of the follow-up period (CD4 count>350 x mm3).The variables HIV stage at diagnosis (X2 556.626; p<0.001); viral load at the start of antiretroviral therapy >100,000 cop/ml (OR 0.582 95% CI 0.483-0.702; p<0.001); active tuberculosis at diagnosis (OR 0.288 95% CI 0.129-0.645; p=0.001); the inclusion of an integrase inhibitor in the initial antiretroviral therapy (OR 0.601 95% CI 0.481-0.751; p<0.001); changes in antiretroviral therapy (OR 1.794 95%CI 1.432-2.249; p<0.001); adherence to antiretroviral therapy dispensing above 95% (OR 1.534 95% CI 1.208-1.946; p<0.001); the record of >3 consultations by the I.D specialist in the period of follow-up (OR 1.539 IC95% 1.194-1.984; p<0.001) met the criterion of significance. (See table No. 3).

The performance for immunological reconstitution of the multivariable logistic regression model obtained an AUC/ROC of 0,8364, showing better predictive accuracy than for the other previous outcomes (See figure 3).

**Figure 3.**
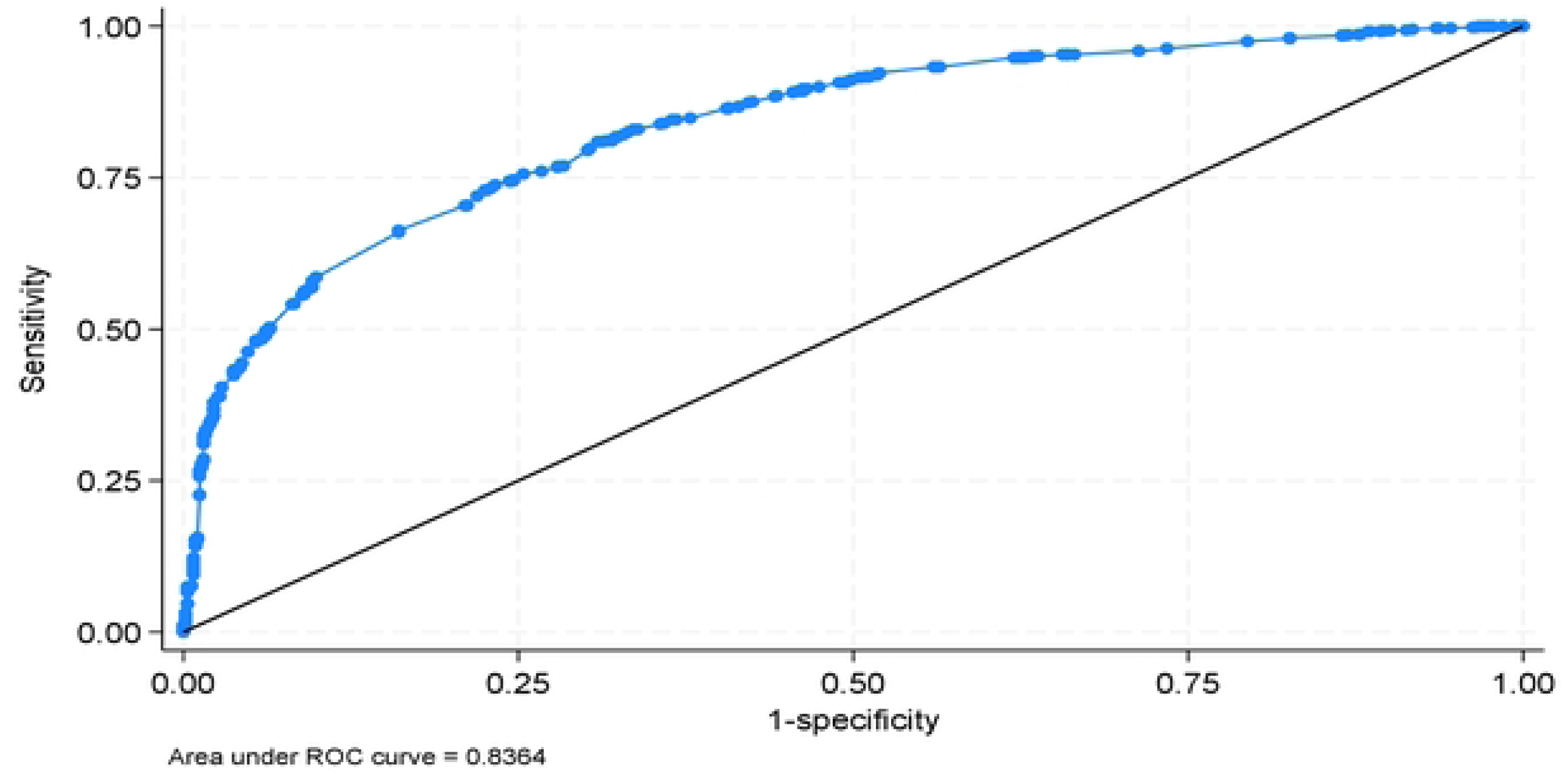
AUC/ROC for “immunological reconstitution” multivariable logistic regression.

For the artificial neural network, the model for the “suppressed viral load” outcome achieved an overall correct percentage in the training phase of 84.5% and in the tests of 84% (See figure 4). An AUC/ROC of 0.777 for this outcome. In this same model it showed 15.5% of incorrect forecasts in the training phase and 16% in the tests. The relationships of the neural network with its synaptic weighting can be observed in figure 4. Evaluating against its predictive capacity, this neural network showed an accuracy of 80.9% and a Kappa value of 67.3%.

**Figure 4.**
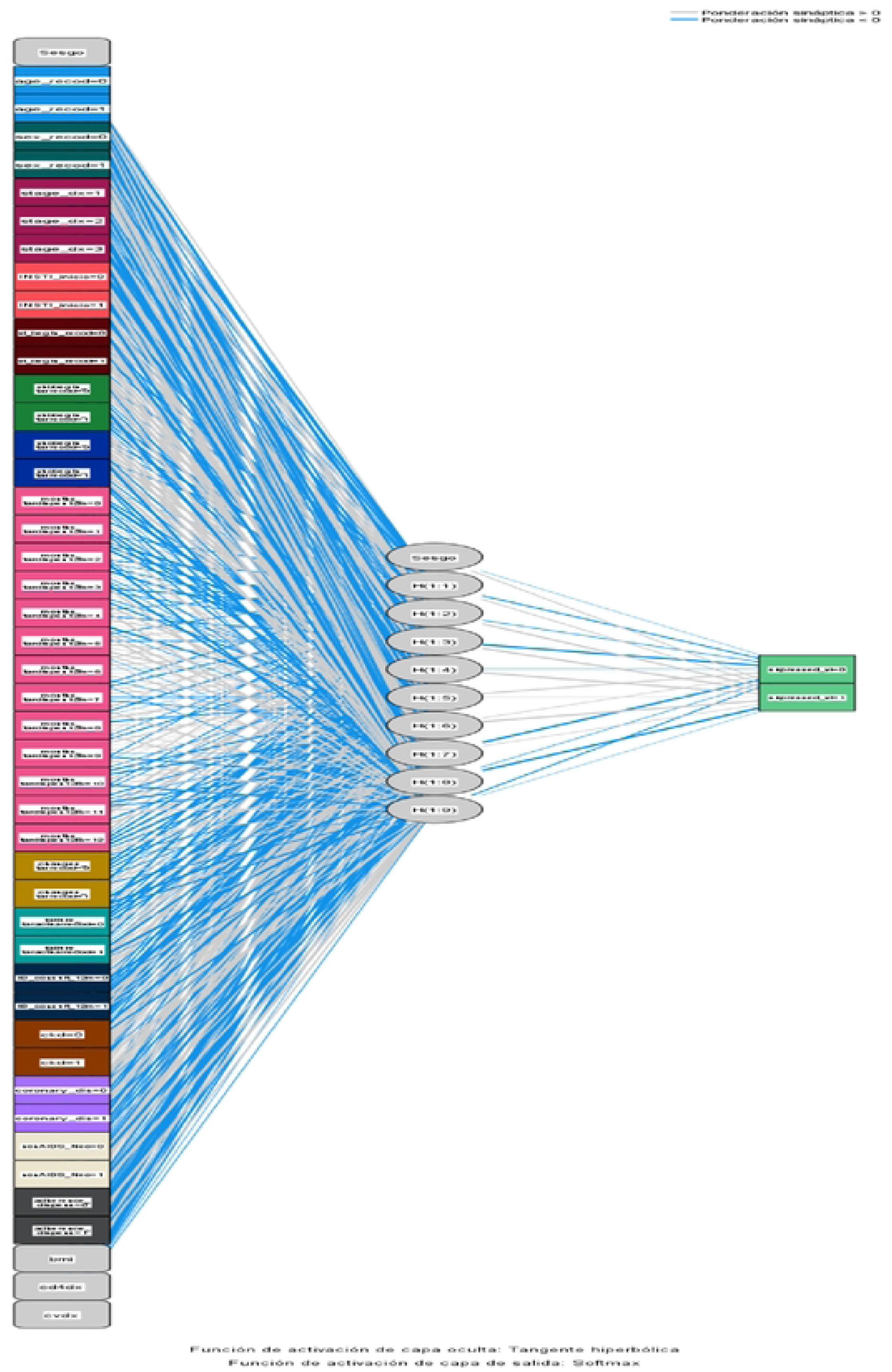
Artificial neural network for suppressed viral load.

The accuracy of the neural network obtained through the AUC/ROC curve was 0.777 for this outcome.

In predicting the outcome of undetectable viral load, the neural network obtained an accuracy through the evaluation of AUC/ROC of 0.69, reflecting a similar performance when we compare it with the logistic regression model, and being consistently inferior for both models when compared with de suppressed viral load outcome. (See figure 5).

**Figure 5.**
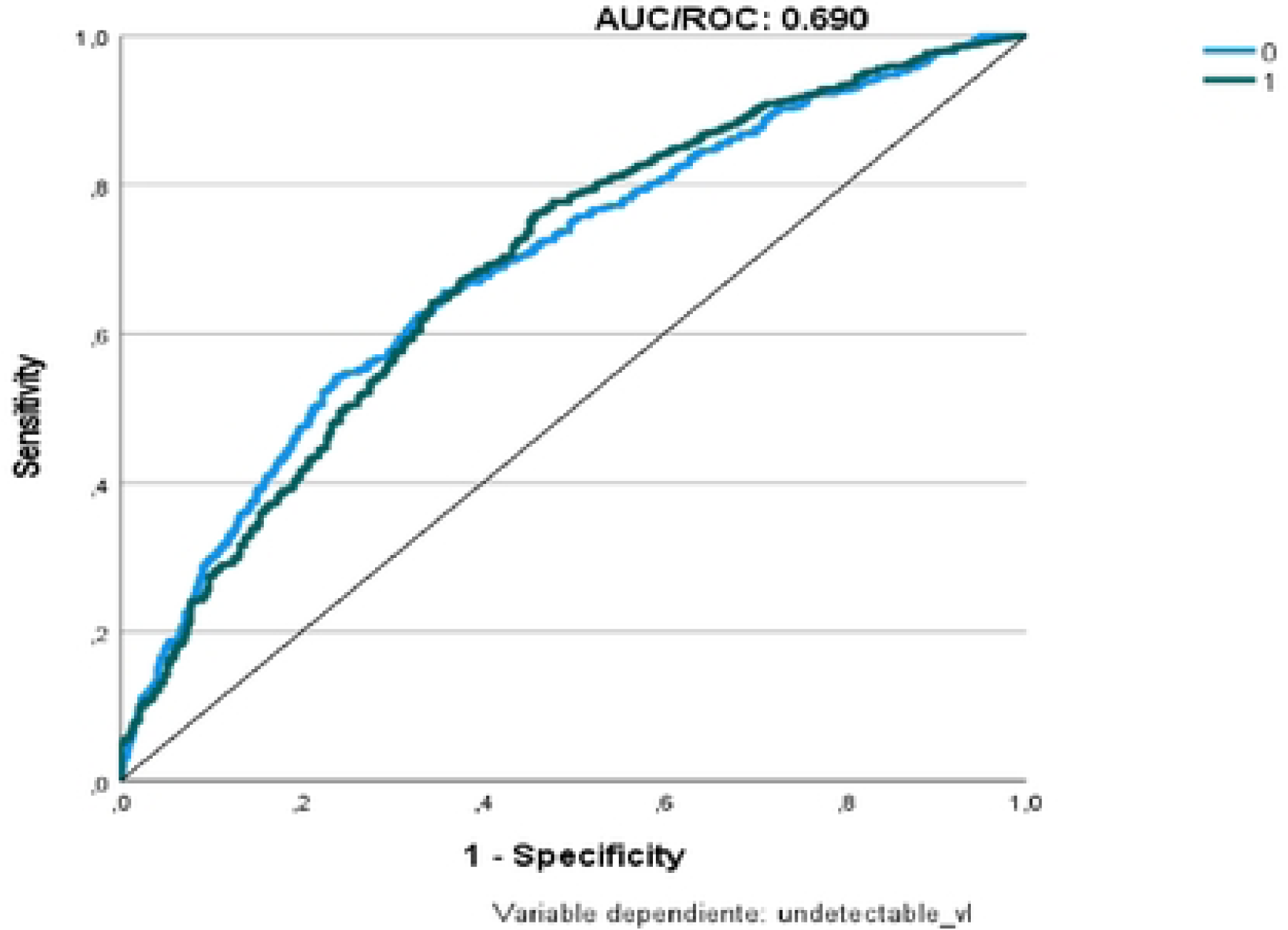
AUC/ROC curve for “undetectable viral load” ANN.

The best performance observed in the neural network models was observed for the outcome of immunological reconstitution, obtaining an AUC/ROC of 0.878, consistent with the same finding for the performance of the multivariable logistic regression models. (See figure 6).

**Figure 6.**
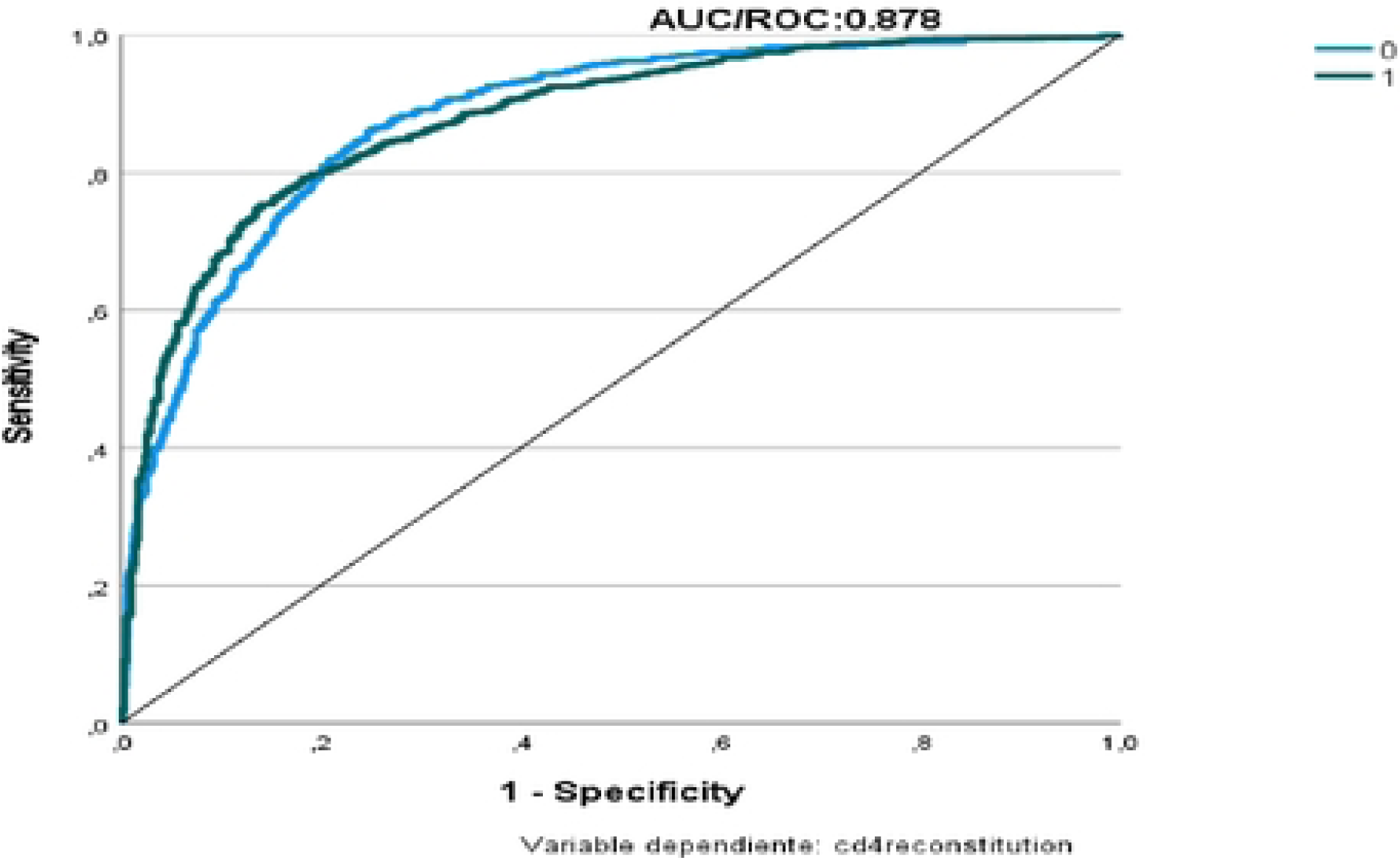
AUC/ROC curve for “immunological reconstitution” ANN.

When we compared the results of both modelling strategies for the three mentioned outcomes we see similar performances, favoring by a slight difference the neural network models and being consistent in better results for the prediction of immunological reconstitution in both cases. (See table 4).

**Table 4.** AUC/ROC Comparison Logistic regression models vs multilayer perceptron neural networks.

| Predictive models | AUC/ROC |
| --- | --- |
| Multivariate logistic regression |  |
| Suppressed viral load | 0,70 |
| Undetectable viral load | 0,66 |
| Immunological reconstitution | 0,83 |

| Multilayer Neural Network/Perceptron |  |
| --- | --- |
| Suppressed viral load | 0,77 |
| Undetectable viral load | 0,69 |
| Immunological reconstitution | 0,87 |

## Discussion

In this study we applied predictive models using the multivariable logistic regression and artificial neural networks for three predefined outcomes related to virological and immunological success in the treatment of people living with HIV (PLWH), the variables included those routinely collected through the SISCAC database in Colombia, including demographic information, clinical characteristics, and laboratory parameters. The SISCAC database purpose is to centralize the registration and validation in real time of quality and assistance indicators from patients of the high-cost account of the Colombian health system (https://cuentadealtocosto.org/general/comunicado-siscac/).

We conducted a statistical analysis with a predictive intention, mainly referencing in the study by Wang et al, who developed machine learning algorithms through artificial neural networks, random forests, and support vector machines, to predict viral load results from 1204 episodes of change in antiretroviral therapy; incorporating genotype mutations input and treatment history variables (17).

Our population consisted of mainly men (86%), observing 75% of all below de 42 years of age, and with a fairly good level for CD4 lymphocyte count both at diagnosis and the initiation of antiretroviral therapy; suggesting earlier introductions of antiretroviral therapy for this cohort. We found previously known variables predictive of success such as the stage for HIV/AIDS at diagnosis, the presence of therapy failures during follow-up and the proportion of adherence to overall antiretroviral dispensation by the provider of care. No association was found both for the suppressed viral load and undetectable viral load outcomes regarding the initial therapy with the inclusion of an INSTI. An interesting finding was the association of more than three consultations with the I.D specialist during the follow-up period of twelve months with better results in all three outcomes.

A limitation already established neural networks is the necessity of a great amount of data for the training, as much as flexibility is required from the neural network bigger sets of data are needed.

The advantages of neural networks compared to other deep learning or machine learning techniques are:

- Non-linear system: By being able to process information in a non-linear way, it allows the processing of more “chaotic” data.
- Fault tolerance: Since it processes the information in parallel between the different nodes, neural networks are quite tolerant to single or multiple node failures.
- Adaptability: A neural network could adjust its parameters in real time depending on the inputs it receives and its characteristics, while other models do not have this adaptability.

Another weakness of neuronal networks is that once the layers increase, the interpretation of the model becomes more difficult, until it becomes a true black box. In addition, gradients that explode (that is, they grow without limit) or that vanish (that is, they progressively approach zero), constitute aspects that make it difficult to train this architecture and reduce its ability to process long-duration sequences.

Although neural networks significantly reduce this inconvenience, they do not eliminate it and this problem becomes more evident as the sequence to process becomes increasingly extensive. The presence of exploding and fading gradients limits the long-term memory of these architectures.

Finally, regarding the results obtained with neural networks, the three models present acceptable results, with high precision and moderate Kappa values. The positive points of the networks neural networks is that they are very flexible and can work with very complex data to find patterns that other techniques cannot; and while in the other techniques you have more or less information about the patterns found in the data and how the variables affect the result, with neural networks this information is lost and you have only one input and one output.

Regarding the comparison of the models, we found similar performance both for the multivariable logistic regression and the neural networks, in both cases the accuracy was superior in predicting the immunological reconstitution outcome, and in this matter the inclusion of an INSTI for the initial antiretroviral therapy was statistically significant.

Also, we must consider the risk for selection bias, given that we obtained data from only one HIV care provider in the country, which can affect external validity, but as this institution allowed access to data from patients from three different cities in different circumstances of health insurance, access to care and socioeconomical conditions, the bias can be minimized.

## Conclusions

This is an important exercise from already verified data uploaded to a nationwide database, allowing access to several variables from people living with HIV (PLWH) in Colombia; it allows an evolving process of integrating artificial intelligence algorithms to provide a growing framework directed to support health care providers with new tools towards their patients.

## Competing interests

The authors of this manuscript declare they have no potential conflicts of interest with respect to the research, authorship, and/or publication of this article.

## Ethical responsibilities

The authors manifest that this study followed the recommendations for the secondary use of clinical data (24–26). The protocol was reviewed and approved by the Vidamedical IPS research and ethics committee in the corresponding session of march 17^th^, 2023.

## Data Availability

The data that support the findings of this study are available on request from the corresponding author. The data are not publicly available due to privacy or ethical restrictions.

https://cuentadealtocosto.org/site/general/comunicado-siscac/.

## Acknowledgements

We thank Vidamedical IPS (https://www.vidamedicalips.com/) for supporting this study at its HIV Clinics in Colombia

## Funding

This work was executed with funding from the investigators, personal computing equipment and statistical programs with individual licensing from them. No external nor sponsor funding was used.

## List of abbreviations

AIDS: Acquired immune deficiency syndrome
ANN: Artificial neural network
ART: antiretroviral therapy
I.D: Infectious Diseases
INSTI: Integrase strand transfer inhibitors
PLWH: people living with HIV
S.D: Standard deviation

## Notes

### Competing Interest Statement

The authors have declared that no competing interests exist.

### Funding Statement

The author(s) received no specific funding for this work.

### Author Declarations

* This research work was approved by VIDAMEDICAL's research ethics committee. * The project involves research on human subjects and complies with the Scientific, Technical and Administrative Standards for Health Research established in Resolution No. 008430 of 1993 and Resolution 2378 of 2008. * Informed consent was not required since databases were reviewed and analyzed anonymously.

